# Sex–associated differences between body mass index and SARS-CoV-2 antibody titers following the BNT162b2 vaccine among 2,435 healthcare workers in Japan

**DOI:** 10.1101/2021.08.30.21262862

**Authors:** Shohei Yamamoto, Tetsuya Mizoue, Akihito Tanaka, Yusuke Oshiro, Natsumi Inamura, Maki Konishi, Mitsuru Ozeki, Kengo Miyo, Wataru Sugiura, Haruhito Sugiyama, Norio Ohmagari

**Affiliations:** Department of Epidemiology and Prevention, Center for Clinical Sciences, National Center for Global Health and Medicine, Tokyo, Japan; Department of Laboratory Testing, Center Hospital of the National Center for the Global Health and Medicine, Tokyo, Japan; Center for Medical Informatics Intelligence, National Center for Global Health and Medicine, Tokyo, Japan; Center for Clinical Sciences, National Center for Global Health and Medicine, Tokyo, Japan; Center Hospital of the National Center for the Global Health and Medicine, Tokyo, Japan; Disease Control and Prevention Center, National Center for Global Health and Medicine, Tokyo, Japan

**Keywords:** Body mass index, Obesity, SARS-CoV-2, Vaccine, Immune response

## Abstract

Obesity may downregulate vaccine-induced immunogenicity, but the epidemiological evidence for the COVID-19 vaccine is limited, and the sex-associated difference is unknown. It was observed that a higher body mass index was associated with lower titers of spike IgG antibodies against SARS-CoV-2 in men but not in women.

## Introduction

Obesity is a major risk factor for morbidity and mortality in patients with coronavirus disease 2019 (COVID-19) [1]. Given the detrimental effect of excess adipose tissue on the immune system [2], obesity may interfere with the production of vaccine-specific antibodies. In clinical trials on the mRNA-based vaccine against severe acute respiratory syndrome coronavirus (SARS-CoV-2), vaccine recipients had a markedly lower risk of COVID-19 irrespective of obesity status [3]. However, more recent observational studies have shown that the efficacy of the vaccine appears to attenuate over time, which is concerning, especially after the rapid spread of the Delta variant [4]. In this context, whether and how obesity influences the immunogenicity of the COVID-19 vaccine remains unanswered.

The current evidence regarding the association between BMI and vaccine-induced antibody titers is limited and inconsistent [5-7]. Given the sex differences in the distribution and function of adipocytes [8], the inconsistency among previous studies could be ascribed, at least in part, to the lack of consideration of the sex. Here, we investigated the sex-associated difference between body mass index (BMI) and SARS-CoV-2 antibodies among 2,435 healthcare workers who received two doses of the BNT162b2 vaccine.

## Methods

We conducted a serological survey in June 2021 among workers of the National Center for Global Health and Medicine, Japan (NCGM), where a vaccination program (COVID-19 mRNA-LNP BNT162b2; Pfizer-BioNTech) was conducted between March and June 2021. Of the 3,092 workers invited, 2,763 (89%) participated. In the present study, we included those who had received two doses of the vaccine (n=2,514) but excluded those who attended the survey within 14 days of the second vaccination (n=5) and those who lacked data on height or weight (n=74), leaving 2,435 for analysis. Written informed consent was obtained from each participant, and the study procedure was approved by the NCGM ethics committee.

Participants were asked to donate venous blood. Data on height, weight, vaccination history (confirmed against records), and history of COVID-19 were collected through a questionnaire. BMI was calculated as the weight (kg) divided by the square of the height (m) and participants were grouped into six categories, ranging from <18.5 kg/m^2^ to ≥27 kg/m^2^. IgG against the SARS-CoV-2 spike protein was quantitatively measured (AdviseDx SARS-CoV-2 IgG II assay, Abbott ARCHITECT®; positive threshold: ≥50.0 AU/mL). Linear regression modeling was used to estimate the means of log_10_-transformed IgG titers for each sex and BMI category while adjusting for age (continuous), days after the second vaccination (continuous), and history of COVID-19, and the values obtained were then back-transformed to present geometric means. The interaction between sex and trend was also tested. Statistical significance was set at P<0.05 for trend and P<0.1 for interaction tests. Statistical tests were conducted using Stata 17.0.

## Results

The mean age was 38.5 years [interquartile range (IQR): 27.6–47.6], 70% were women, 0.57% had a history of COVID-19, and only 2.7% were obese (BMI ≥30 kg/m^2^). The median interval between the second vaccination and blood sampling was 64 days (range, 15–103 days). All participants were positive for SARS-CoV-2 spike antibodies after the assay, with a median of 6,063 AU/mL (IQR: 3,620–9,919).

There was a significant interaction between the BMI category and sex (P=0.07; **Figure 1**). In men, SARS-CoV-2 spike antibody titers progressively decreased with increasing BMI (P for trend <0.01); adjusted geometric means [95% CIs] were 6,096 [4,876–7,621] and 4,615 [4,063–5,242] for BMI categories <18.5 kg/m^2^ and ≥27 kg/m^2^, respectively. In women, there was no significant difference in antibody titers across BMI categories (P for trend = 0.46); adjusted geometric means [95% CIs] were 6176 [5,718–6,670] and 6,158 [5,396–7,027] for BMI categories <18.5 kg/m^2^ and ≥27 kg/m^2^, respectively. Men with a BMI of <18.5 (P=0.91) had comparable antibody titers to women in the same BMI category, whereas men in the remaining BMI categories from 18.5–20.9 kg/m^2^ to ≥27 kg/m^2^ had significantly lower antibody titers than women in the same category (P<0.05), and the sex-associated difference appeared to be more pronounced with increasing BMI.

**Figure 1.**
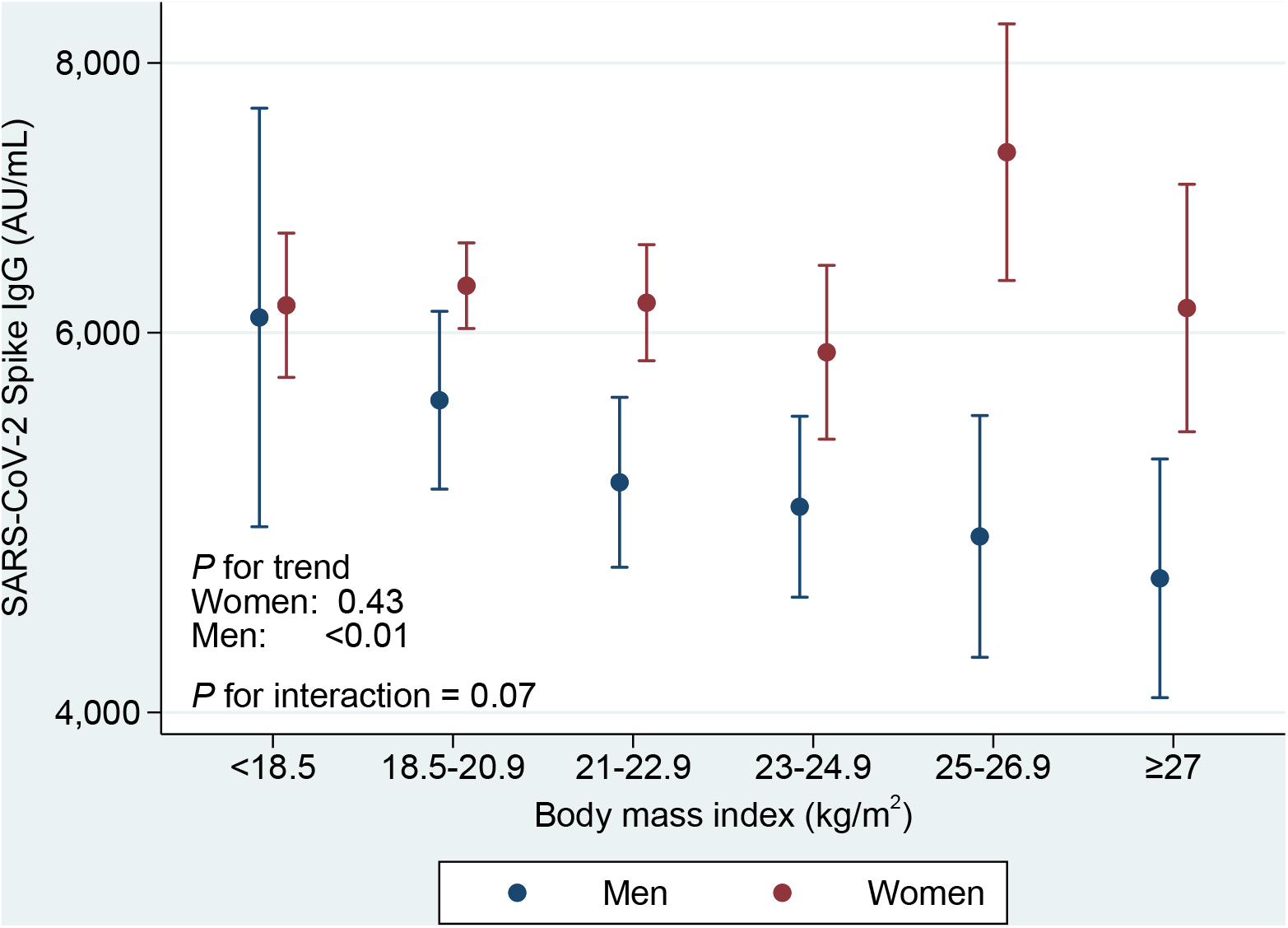
Estimated geometric means of SARS-CoV-2 spike IgG titers with 95 % CIs by BMI and sex after the second vaccination dose. Data are shown as geometric means with 95 % CIs estimated by linear regression model adjusting for age (continuous), days after the second vaccination (continuous), and history of COVID-19. The number of participants in the categories of <18.5, 18.5–20.9, 21–22.9, 23–24.9, 25–26.9, and ≥27 kg/m^2^ were 256, 721, 390, 174, 80, and 86 for women, and 30, 168, 182, 164, 91, and 93 for men, respectively. The adjusted geometric means [95% CIs] for lowest to highest BMI categories were 6096 [4876–7621], 5581 [5076–6136], 5113 [4670–5598], 4981 [4523–5486], 4826 [4242–5490], and 4615 [4063–5242] in men; 6176 [5718–6670], 6306 [6025–6601], 6193 [5821–6588], 5874 [5353–6445], 7272 [6341–8339], and 6158 [5396–7027] in women, respectively. BMI: body mass index, CIs: confidence interval, SARS-CoV-2: severe acute respiratory syndrome coronavirus.

## Discussion

In this cross-sectional study among healthcare workers who received two doses of the BNT162b2 vaccine (median interval between the second vaccination and blood draw of 64 days), all participants, irrespective of BMI, were seropositive, with most participants having high titers of SARS-CoV-2 spike IgG antibodies, which is compatible with the results of clinical trials (median follow-up of 2 months), showing no difference in effectiveness between obese and non-obese vaccine recipients [3].

Nevertheless, we found a marked sex-associated difference between BMI and SARS-CoV-2 spike IgG antibody titers, with a significant inverse association in men but not in women. To the best of our knowledge, this is the first report to specifically address the differential role of sex in the association between adiposity and SARS-CoV-2 vaccine-induced immunogenicity.

One possible mechanism behind the lower antibody titers associated with higher BMI in men is the dysregulated production of adipocytokines in obesity. With increasing adipose tissue, leptin, TNF-α, and IL-6 are overproduced, while adiponectin decreases [2] Adipocytokine profile imbalance can cause low-grade chronic inflammation, which can induce B cell immunosenescence and interfere with antibody production post-vaccination [9].

On the other hand, antibody titers did not change with increasing BMI in women. Given that 82% of female participants were under the age of 50 in the present study, the sex-associated difference could be ascribed to the effect of sex hormones (estrogen, testosterone, and progesterone) on the regulation of adipose tissue distribution, adipogenesis, and adipocyte metabolism and production [8]. Owing to these hormones, women have less visceral adipose tissue, greater brown adipose tissue, and higher circulating adiponectin concentrations compared to men [8], all of which have been suggested to suppress chronic inflammation and activate the immune system [2, 10].

In summary, higher BMI in men, but not in women, was associated with a lower antibody titer after vaccination, suggesting the need for attentive monitoring of vaccine efficacy over time in obese men, who are also at greater risk of higher severity and morbidity from COVID-19 infection. We hope that the present findings inform the decisions of policymakers and healthcare professionals regarding the selection of persons of priority for the administration of the booster vaccination, which has already begun in certain countries.

## Data Availability

The datasets generated and/or analyzed during the current study are not publicly available due to ethical restrictions and participant confidentiality concerns, but de-identified data are available from the corresponding author to qualified researchers on reasonable request.

## Acknowledgements

We thank Haruka Osawa for her contribution to data collection.

## References

1. Popkin BM, D. S, Green WD, et al. Individuals with obesity and COVID-19: A global perspective on the epidemiology and biological relationships. Obes Rev 2020; 21(11).

2. De Heredia FP, Gómez-Martínez S, Marcos A. Obesity, inflammation and the immune system. Proc Nutr Soc 2012; 71(2): 332–8.

3. Polack FP, Thomas SJ, Kitchin N, et al. Safety and Efficacy of the BNT162b2 mRNA Covid-19 Vaccine. N Engl J Med 2020; 383(27): 2603–15.

4. Pouwels, K. B. et al. Preprint at Univ. Oxford https://www.ndm.ox.ac.uk/files/coronavirus/covid-19-infection-survey/finalfinalcombinedve20210816.pdf (2021).

5. Mitsunaga T, Ohtaki Y, Seki Y, et al. The evaluation of factors affecting antibody response after administration of the BNT162b2 vaccine: A prospective study in Japan. medRxiv 2021: 2021.06.20.21259177.

6. Pellini R, Venuti A, Pimpinelli F, et al. Initial observations on age, gender, BMI and hypertension in antibody responses to SARS-CoV-2 BNT162b2 vaccine. EClinicalMedicine 2021; 36: 100928.

7. Bayart J-L, Morimont L, Closset M, et al. Confounding Factors Influencing the Kinetics and Magnitude of Serological Response Following Administration of BNT162b2. Microorganisms 2021; 9(6): 1340.

8. Karastergiou K, Smith SR, Greenberg AS, Fried SK. Sex differences in human adipose tissues – the biology of pear shape. Biol Sex Differ 2012; 3(1): 13.

9. Frasca D, Blomberg BB. Adipose Tissue Inflammation Induces B Cell Inflammation and Decreases B Cell Function in Aging. Front Immunol 2017; 8: 1003.

10. Peterson KR, Flaherty DK, Hasty AH. Obesity Alters B Cell and Macrophage Populations in Brown Adipose Tissue. Obesity 2017; 25(11): 1881–4.

